# Trends, Outcomes, and Healthcare Disparities of Asian patients with Heart Failure and Advanced Chronic Kidney Disease

**DOI:** 10.1101/2023.10.09.23296780

**Authors:** Samuel S. Tan, Wenchy YY. Tan, Lucy S. Zheng, Shasawat Kumar

**Affiliations:** Department of Medicine, Mount Sinai Beth Israel, New York, New York, USA; Department of Population Health Sciences, Weill Cornell, New York, New York, USA

## Abstract

**Background:** Data on outcomes and risk factors for Asian patients with advanced chronic kidney disease admitted for heart failure are limited. The purpose of this study was to investigate contemporary racial and socioeconomic disparities in Asian patients with advanced kidney disease admitted for heart failure exacerbation.

**Methods:** This was a retrospective cohort study that utilized data from the National Inpatient Sample between January 2016 and December 2020. Patients who had a primary diagnosis of HF and a concomitant diagnosis of advanced CKD were included. The primary outcome of interest was in-hospital mortality. Secondary outcomes include hospital cost, length of stay, and other clinical outcomes. Weighted multivariable logistic regression was used to adjust for comorbidities.

**Results:** A total of 186,431 admissions for heart failure exacerbation with advanced kidney disease were identified, of which 10.7% (*n =* 20,006) were of Asian descent. Asian patients had the lowest prevalence of comorbidities compared with all other ethnicities. After adjusting for comorbidities, sex, and age, Whites (OR: 1.11; 95% CI 1.03 to 1.20; p<0.009) had higher odds of mortality, while Black (OR: 0.58; 95% CI 0.53 to 0.63; p< 0.001) and Hispanic patients (OR: 0.69; 95% CI 0.62 to 0.78; p< 0.001) had lower odds of mortality compared to Asian patients.

**Conclusion:** Despite recent efforts to address healthcare inequality, health disparities among Asian patients with HF and CKD remain. The significant difference in odds of mortality and complications, despite a lower comorbidity burden, indicates the importance of further research regarding the possible causality of such disparities.

## Introduction

Heart failure (HF) is an increasingly common cause of hospitalization in the US and affects over 6.2 million individuals in the United States alone.^1^ From 2014 to 2017, heart failure hospitalization increased from 2.4 to 4.9 per 1000 persons.^2^ Despite recent advances in pharmacologic therapy, prevalence is estimated to increase by 46% by 2030. A systematic review estimated that the annual median medical cost for heart failure specific hospitalization is $15,879 per year^3^ while US heart failure costs are expected to be at least $70 billion per year, with a total cost of caring for patients with heart failure reaching $160 billion by 2030.^4^

Chronic Kidney Disease (CKD) impacts 37 million Americans, 15% of the adult population or approximately one in seven Americans.^5^ Socioeconomic inequalities in cardiovascular and renal diseases present major and persistent global public health challenges. Previous research has indicated a higher prevalence of HF associated with CKD was in the African American and Hispanic populations, in addition to disparity in the prescription rates of innovative medicines between racial and ethnic minorities and individuals of White ethnicity.^6^ HF and CKD are major public health issues with gender, race, ethnicity, and socioeconomic level differences in incidence, prevalence, and complications. One study showed that racial disparities have increased healthcare expenditures by $230 billion and indirect costs from premature illness and death by $1 trillion.^7^

Collectively, Asian’s are the world’s largest population although most HF data comes from Europe and North America.^8^ Data on risk factors and outcomes for Asian patients admitted for HF exacerbation are limited, and those with concomitant advanced CKD are even less well understood. To address this knowledge gap, we used a nationally representative database, ensuring a sizable study population that enhances generalization of results. We also examined sex, race, and ethnicity disparities in in-hospital all-cause mortality, concomitant risk factors, and comorbidities, as well as 5-year temporal trends.

## Methods

### Data sharing statement

The National Inpatient Sample (NIS) data set used in this project is publicly available.

### Data source

The study utilized data from the National Inpatient Sample (NIS) from January 1, 2016, to December 31, 2020. The NIS is a comprehensive all-payer database for inpatient treatment in the United States by the Healthcare Cost and Utilization Project (HCUP), a component of the Agency for Healthcare Research and Quality (AHRQ). With over seven million hospital admissions, the dataset covers 97% of the U.S. population and a 20% stratified sample of community hospitals. International Classification of Diseases-10th Edition-Clinical Modification (ICD-10-CM) and International Classification of Diseases-10th Edition-Procedural Coding System (ICD-10-PCS) codes are used for up to 38 diagnoses and 15 procedures related to that stay. Median household annual income was categorized into 4 quartiles: 0 to the 25th percentile quartile ($1–$49,999), 26th to the 50th quartile ($50,000–$64,999), 51st to the 75th quartile ($65,000–$85,999), and 76th to the 100th quartile ($86,000+). The Core-Based Statistical Area (CBSA) criteria determined whether a hospital was urban or rural. Because we used publicly available data with no patient identifiers, this study was exempt from formal review by the Institutional Review Board of our institution.

### Patient selection

We used ICD-10-CM I50.x and I11.0 to identify adult patients aged 18 and over who had a primary diagnosis of heart failure. ICD-10-CM codes I13.0, I13.2, N18.4, N18.5, N19.6, N18.9, and Z99.2 were used to identify those with advanced CKD (**Supplemental Table 1**). Patients were then categorized by race and ethnicity (Asian, White, Black, Hispanic). All diagnosis fields were accounted for to identify the study population. The study overview and methods flowsheet are shown in **Figure 1**.

**Figure 1.**
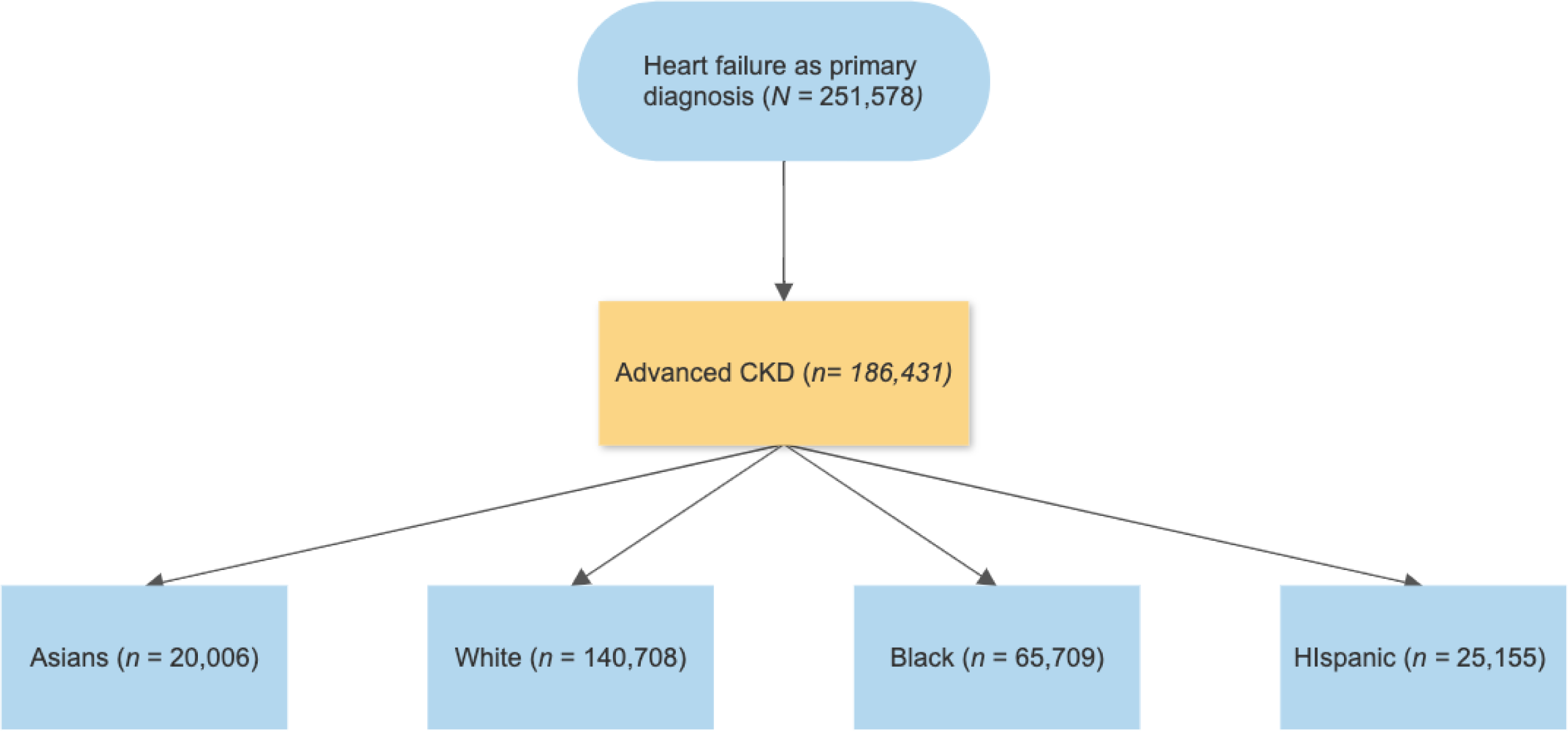
Sampled Population Flow Chart.

### Study population

Patient characteristics (ethnicity, race, age, sex, insurance, income), hospital characteristics (teaching status and location, region, bed size) and Elixhauser comorbidities were identified from the HCUP-NIS database. A list of ICD-10-CM codes used to identify comorbidities and procedural characteristics can be found in **Supplement Table 2**. The Elixhauser comorbidity index was used to identify the burden of comorbid diseases.

### Outcomes

Primary outcome of interest was in-hospital mortality. Secondary outcomes include hospital cost of stay, length of stay, and other clinical outcomes. Clinical outcomes included: acute kidney injury (AKI), acute myocardial infarction (AMI), acute respiratory failure with hypoxia, acute stroke, cardiac tamponade, cardiogenic shock, cardiopulmonary resuscitation, use of mechanical ventilation, use of mechanical circulatory support (ECMO, IABP, pVAD), pulmonary embolism, SIRS, sepsis, ventricular fibrillation, ventricular flutter, ventricular tachycardia.

### Statistics

As recommended by HCUP-NIS, survey procedures using discharge weights provided by HCUP-NIS data were used to generate national estimates. Sample characteristics were summarized using the mean for continuous variables and proportions for categorical variables. To examine the primary and secondary outcomes, we used multinomial propensity score methodology to match hospitalizations with HF exacerbation and advanced chronic kidney disease comparing Asian, White, Black, and Hispanic patients. Variables in the analysis included all baseline comorbidities found in **Table 2**. Multivariable analysis was conducted to identify clinical and hospital characteristics independently associated with HF exacerbation and advanced CKD. The Healthcare Cost and Utilization Project’s hospital-specific cost-to-charge ratios were used to convert total hospital charges to cost estimates. Total prices were adjusted to US dollars in 2020 using the Consumer Price Index inflation calculator given by the US Bureau of Labor Statistics.^9^ STATA 18 (StataCorp LLC, College Station, TX, USA) was utilized for all statistical analysis. A two-tailed p-value of <0.05 was deemed to be statistically significant. As per the HCUP data use agreement, we did not report variables that contained a small number of unweighted hospitalizations (<11) as this could pose risk of person identification or data privacy violation.

## Results

### Hospitalization characteristics of the study population

There were a total of 251,578 hospitalizations between 2016 and 2020 with a primary diagnosis of heart failure. Of those hospitalizations, 186,431 had a diagnosis of advanced CKD. Asian patients accounted for 20,006 (10.7%) of the final population analyzed, compared to 140,708 (75.5%) White, 65,709 (35.3%) Black, and 25,155 (13.5%) Hispanic patients.

### Patient characteristics

In the Asian population, Medicaid accounted for 13.6% of patients insurance, compared to 4.4%, 16.3%, 18.5% in White, Black, and Hispanic patients, respectively. Median household income Specifically, 55.8% of Black patients and 43.6% of Hispanic patients fell into this income bracket, whereas only 28.9% of Asian and 26.6% of White patients were found in the same range. Asian (22.5%) and White (19.5%) patients were found in the highest quartile of median income, as compared to their Black (8%) or Hispanic (10.4%) counterparts. Detailed baseline and hospital characteristics of the sample population are found in **Table 1** and **Table 2**.

**Table 1.**
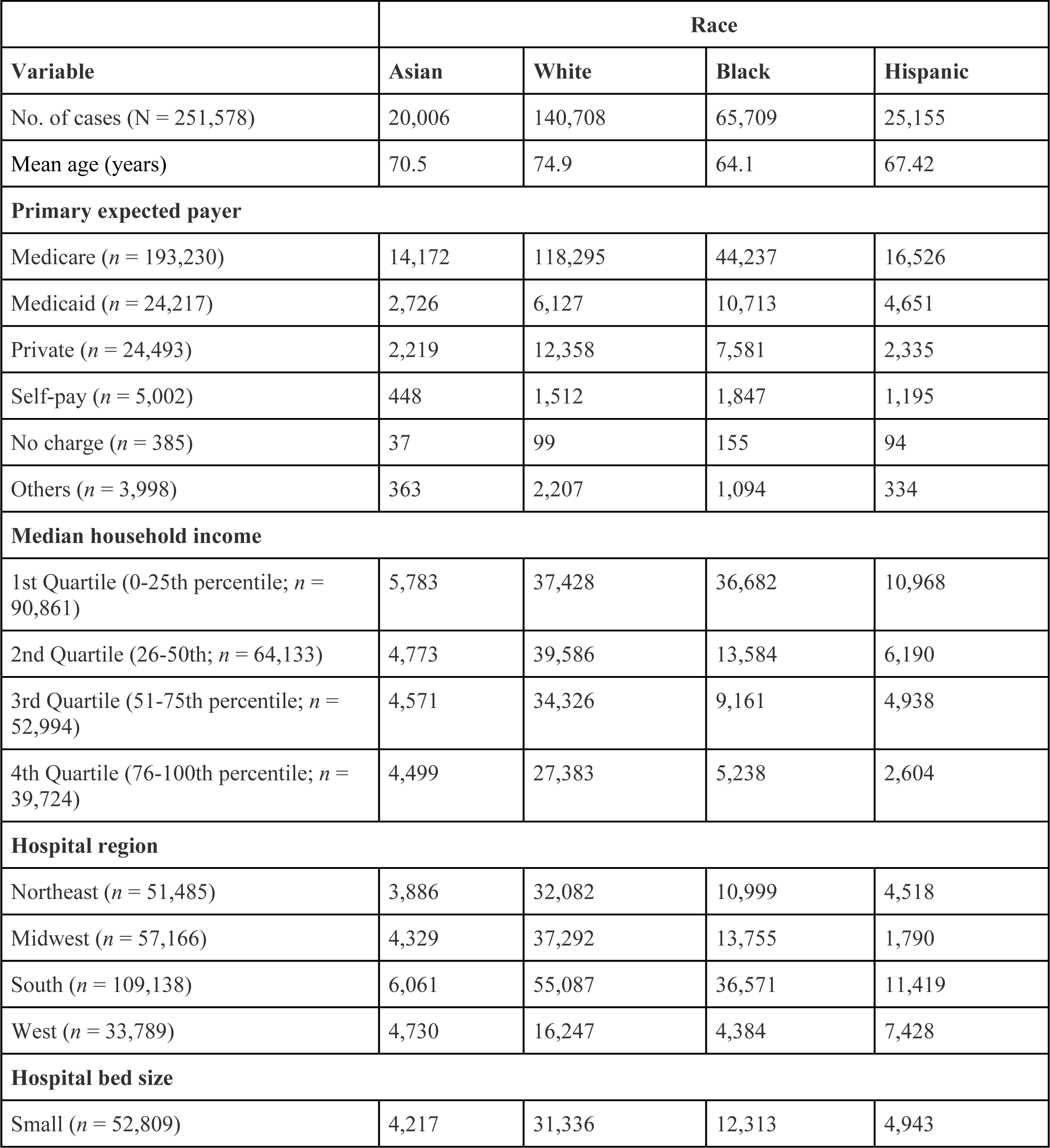
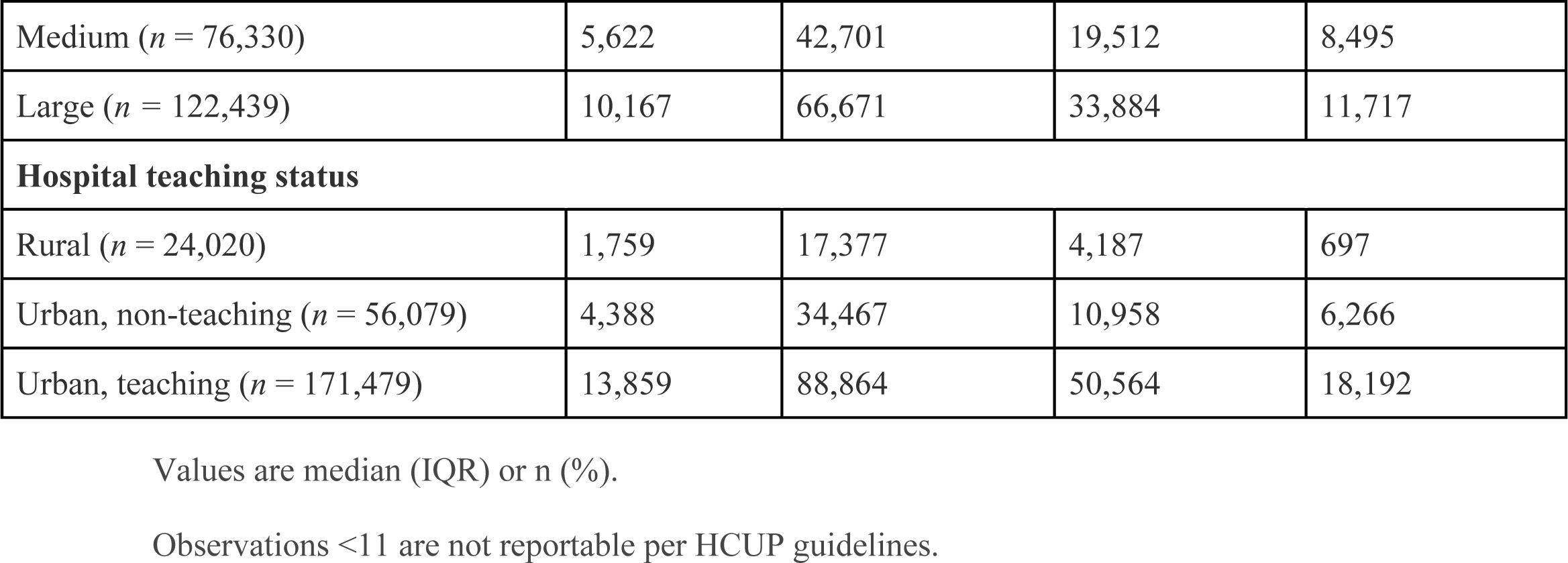
Baseline and Hospital Characteristics of Patients with HF and Advanced CKD.

|  | Race |  |  |  |
| --- | --- | --- | --- | --- |
| Variable | Asian | White | Black | Hispanic |
| No. of cases (N = 251,578) | 20,006 | 140,708 | 65,709 | 25,155 |
| Mean age (years) | 70.5 | 74.9 | 64.1 | 67.42 |
| **Primary expected payer** |  |  |  |  |
| Medicare (*n* = 193,230) | 14,172 | 118,295 | 44,237 | 16,526 |
| Medicaid (*n* = 24,217) | 2,726 | 6,127 | 10,713 | 4,651 |
| Private (*n* = 24,493) | 2,219 | 12,358 | 7,581 | 2,335 |
| Self-pay (*n* = 5,002) | 448 | 1,512 | 1,847 | 1,195 |
| No charge (*n* = 385) | 37 | 99 | 155 | 94 |
| Others (*n* = 3,998) | 363 | 2,207 | 1,094 | 334 |
| **Median household income** |  |  |  |  |
| 1st Quartile (0-25th percentile; *n* = 90,861) | 5,783 | 37,428 | 36,682 | 10,968 |
| 2nd Quartile (26-50th; *n* = 64,133) | 4,773 | 39,586 | 13,584 | 6,190 |
| 3rd Quartile (51-75th percentile; *n* = 52,994) | 4,571 | 34,326 | 9,161 | 4,938 |
| 4th Quartile (76-100th percentile; *n* = 39,724) | 4,499 | 27,383 | 5,238 | 2,604 |
| **Hospital region** |  |  |  |  |
| Northeast (*n* = 51,485) | 3,886 | 32,082 | 10,999 | 4,518 |
| Midwest (*n* = 57,166) | 4,329 | 37,292 | 13,755 | 1,790 |
| South (*n* = 109,138) | 6,061 | 55,087 | 36,571 | 11,419 |
| West (*n* = 33,789) | 4,730 | 16,247 | 4,384 | 7,428 |
| **Hospital bed size** |  |  |  |  |
| Small (*n* = 52,809) | 4,217 | 31,336 | 12,313 | 4,943 |
| Medium (*n* = 76,330) | 5,622 | 42,701 | 19,512 | 8,495 |
| Large (*n* = 122,439) | 10,167 | 66,671 | 33,884 | 11,717 |
| **Hospital teaching status** |  |  |  |  |
| Rural (*n* = 24,020) | 1,759 | 17,377 | 4,187 | 697 |
| Urban, non-teaching (*n* = 56,079) | 4,388 | 34,467 | 10,958 | 6,266 |
| Urban, teaching (*n* = 171,479) | 13,859 | 88,864 | 50,564 | 18,192 |

Values are median (IQR) or n (%).
Observations <11 are not reportable per HCUP guidelines.

**Table 2.**
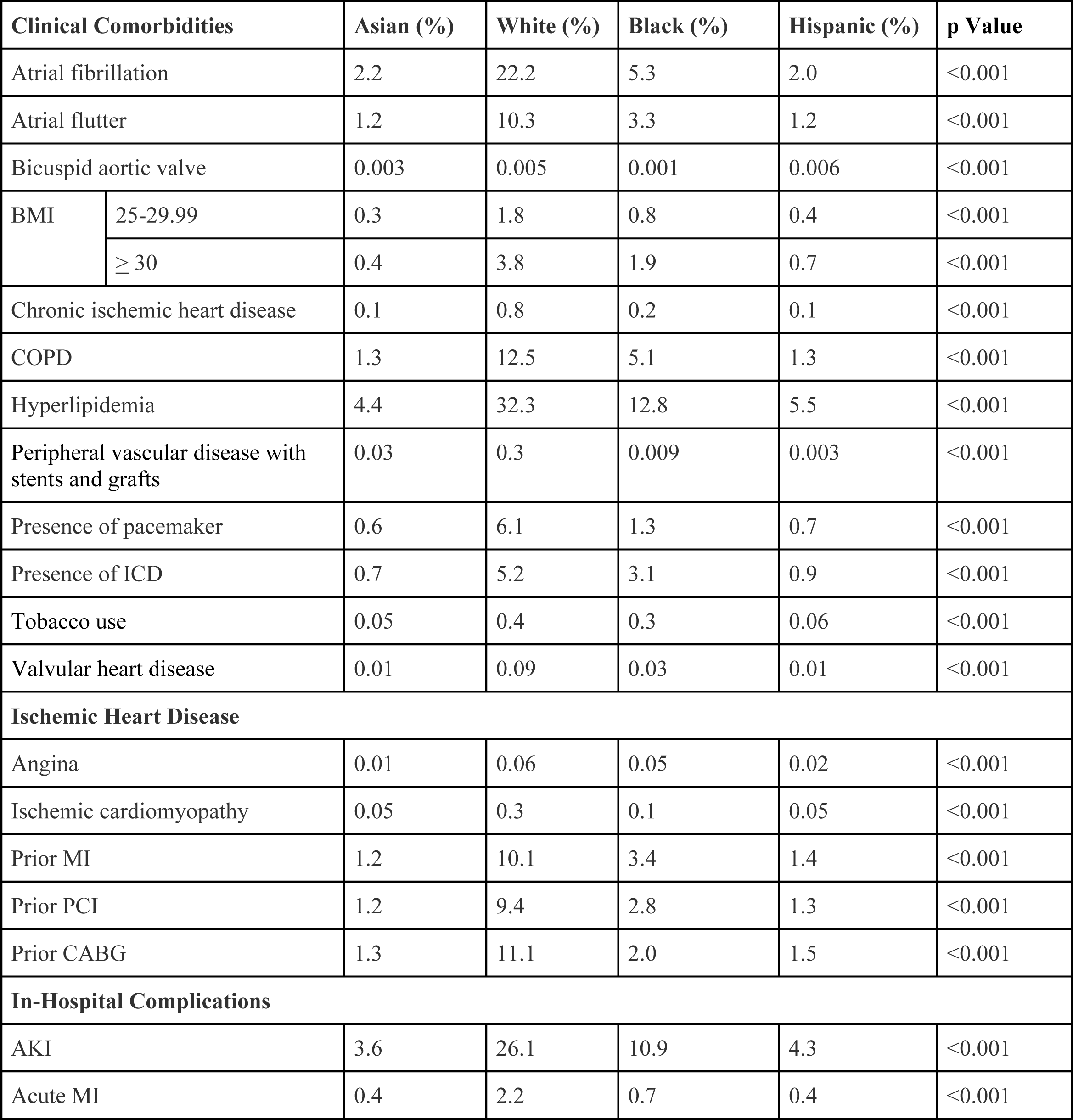
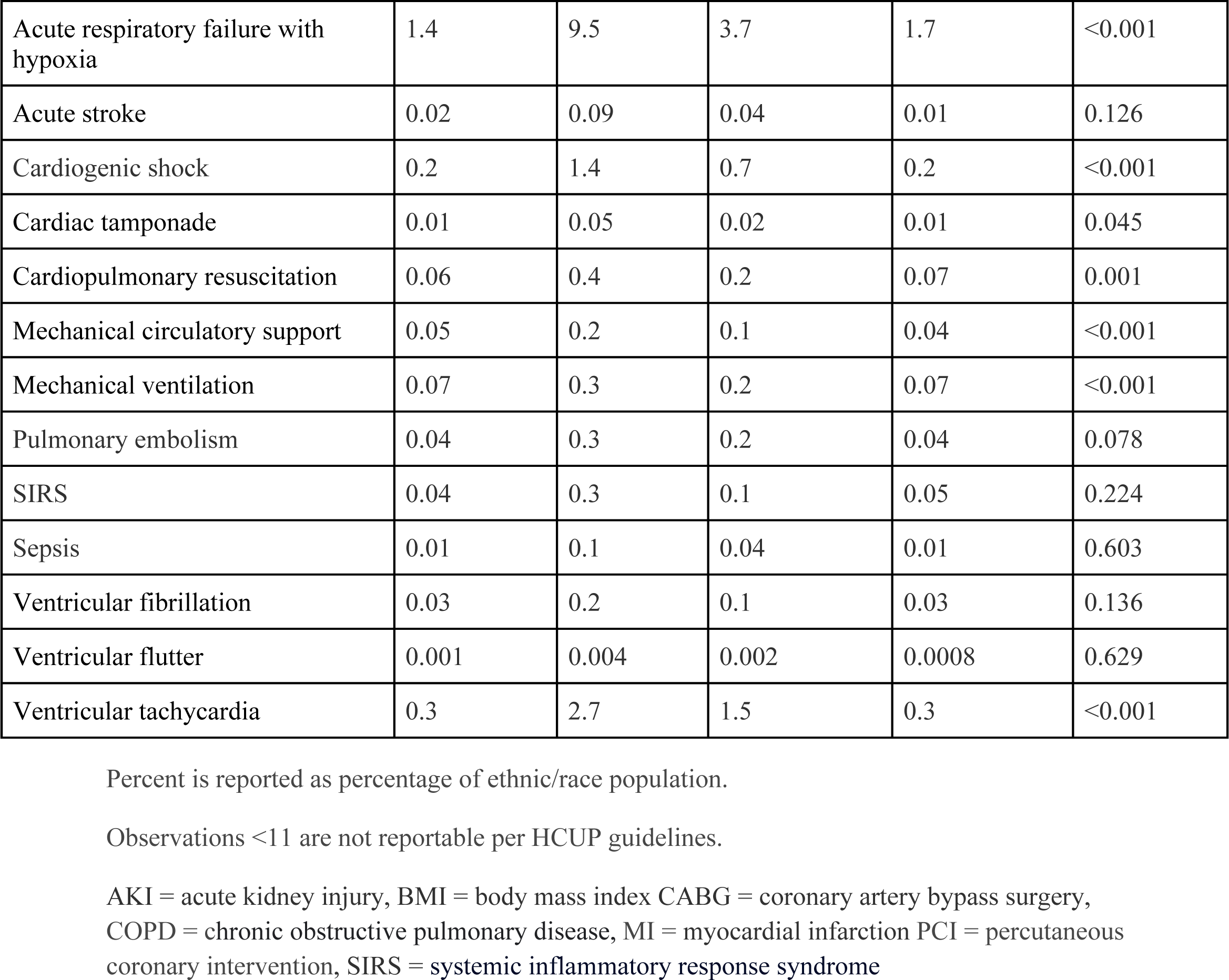
Clinical Comorbidities and In-Hospital Complications of Patients with HF and Advanced CKD.

| Clinical Comorbidities |  | Asian (%) | White (%) | Black (%) | Hispanic (%) | p Value |
| --- | --- | --- | --- | --- | --- | --- |
| Atrial fibrillation |  | 2.2 | 22.2 | 5.3 | 2.0 | <0.001 |
| Atrial flutter |  | 1.2 | 10.3 | 3.3 | 1.2 | <0.001 |
| Bicuspid aortic valve |  | 0.003 | 0.005 | 0.001 | 0.006 | <0.001 |
| BMI | 25-29.99 | 0.3 | 1.8 | 0.8 | 0.4 | <0.001 |
|  | ≥ 30 | 0.4 | 3.8 | 1.9 | 0.7 | <0.001 |
| Chronic ischemic heart disease |  | 0.1 | 0.8 | 0.2 | 0.1 | <0.001 |
| COPD |  | 1.3 | 12.5 | 5.1 | 1.3 | <0.001 |
| Hyperlipidemia |  | 4.4 | 32.3 | 12.8 | 5.5 | <0.001 |
| Peripheral vascular disease with stents and grafts |  | 0.03 | 0.3 | 0.009 | 0.003 | <0.001 |
| Presence of pacemaker |  | 0.6 | 6.1 | 1.3 | 0.7 | <0.001 |
| Presence of ICD |  | 0.7 | 5.2 | 3.1 | 0.9 | <0.001 |
| Tobacco use |  | 0.05 | 0.4 | 0.3 | 0.06 | <0.001 |
| Valvular heart disease |  | 0.01 | 0.09 | 0.03 | 0.01 | <0.001 |
| **Ischemic Heart Disease** |  |  |  |  |  |  |
| Angina |  | 0.01 | 0.06 | 0.05 | 0.02 | <0.001 |
| Ischemic cardiomyopathy |  | 0.05 | 0.3 | 0.1 | 0.05 | <0.001 |
| Prior MI |  | 1.2 | 10.1 | 3.4 | 1.4 | <0.001 |
| Prior PCI |  | 1.2 | 9.4 | 2.8 | 1.3 | <0.001 |
| Prior CABG |  | 1.3 | 11.1 | 2.0 | 1.5 | <0.001 |
| **In-Hospital Complications** |  |  |  |  |  |  |
| AKI |  | 3.6 | 26.1 | 10.9 | 4.3 | <0.001 |
| Acute MI |  | 0.4 | 2.2 | 0.7 | 0.4 | <0.001 |

|  |  |  |  |  |  |
| --- | --- | --- | --- | --- | --- |
| Acute respiratory failure with hypoxia | 1.4 | 9.5 | 3.7 | 1.7 | <0.001 |
| Acute stroke | 0.02 | 0.09 | 0.04 | 0.01 | 0.126 |
| Cardiogenic shock | 0.2 | 1.4 | 0.7 | 0.2 | <0.001 |
| Cardiac tamponade | 0.01 | 0.05 | 0.02 | 0.01 | 0.045 |
| Cardiopulmonary resuscitation | 0.06 | 0.4 | 0.2 | 0.07 | 0.001 |
| Mechanical circulatory support | 0.05 | 0.2 | 0.1 | 0.04 | <0.001 |
| Mechanical ventilation | 0.07 | 0.3 | 0.2 | 0.07 | <0.001 |
| Pulmonary embolism | 0.04 | 0.3 | 0.2 | 0.04 | 0.078 |
| SIRS | 0.04 | 0.3 | 0.1 | 0.05 | 0.224 |
| Sepsis | 0.01 | 0.1 | 0.04 | 0.01 | 0.603 |
| Ventricular fibrillation | 0.03 | 0.2 | 0.1 | 0.03 | 0.136 |
| Ventricular flutter | 0.001 | 0.004 | 0.002 | 0.0008 | 0.629 |
| Ventricular tachycardia | 0.3 | 2.7 | 1.5 | 0.3 | <0.001 |

Percent is reported as percentage of ethnic/race population.
Observations <11 are not reportable per HCUP guidelines.
AKI = acute kidney injury, BMI = body mass index CABG = coronary artery bypass surgery, COPD = chronic obstructive pulmonary disease, MI = myocardial infarction PCI = percutaneous coronary intervention, SIRS = systemic inflammatory response syndrome

All non-Asian patients described as White, Black, and Hispanic respectively had a higher prevalence of comorbidities, notably those associated more disease burden including prior MI (1.2% vs 10.1%, 3.4%, 1.4%), prior PCI (1.2% vs 9.5%, 2.8%, 1.3%), prior CABG (1.3% vs 11.1% vs 2.0% vs 1.5%), presence of pacemaker (0.6% vs 6.1%, 1.3%, 0.7%), and presence of ICD (0.7%, 5.2%, 3.1%, 0.9%) as seem in **Central Illustration.**

### Mortality and hospitalization outcomes

Mortality rate was highest in White (4.2%) patients followed by Asian (3.7%), Black (2.2%), and Hispanic patients (2.6%). White patients were associated with higher odds of mortality compared to Asian patients (OR: 1.11; 95% CI 1.03 to 1.20; p=0.009). Black (OR: 0.58; 95% CI0.53 to 0.63; p< 0.001) and Hispanic patients (OR: 0.69; 95% CI 0.62 to 0.78; p< 0.001) were associated with lower odds of mortality compared to Asian patients.

#### Overall hospitalization outcomes

In comparison to Asian patients, White patients exhibited significantly lower odds of acute MI (OR: 0.88; 95% CI 0.81 to 0.95; p=0.002), cardiogenic shock (OR 0.85; 95% CI 0.77 to 0.95; p=0.003), use of mechanical circulatory support, and mechanical ventilation. Compared to Asian patients, Black patients were associated with lower odds of acute MI (OR: 0.59; 95% CI 0.54 to 0.65; p< 0.001), use of mechanical circulatory support (OR 0.61; 95% CI 0.49 to 0.78; p<0.001), use of mechanical ventilation (OR 0.90; 95% CI 0.80 to 0.98; p<0.001). Hispanic patients were similarly associated with lower odds mechanical circulatory support requirements (OR: 0.60; 95% CI 0.45 to 0.80; p<0.001), mechanical ventilation (OR 0.82; 95% CI 0.71-0.99; p<0.001) and cardiogenic shock (OR: 0.68; 95% CI 0.59 to 0.79; p<0.001). Additional details on mortality and in-hospital complications can be found in **Table 3**.

**Table 3.**
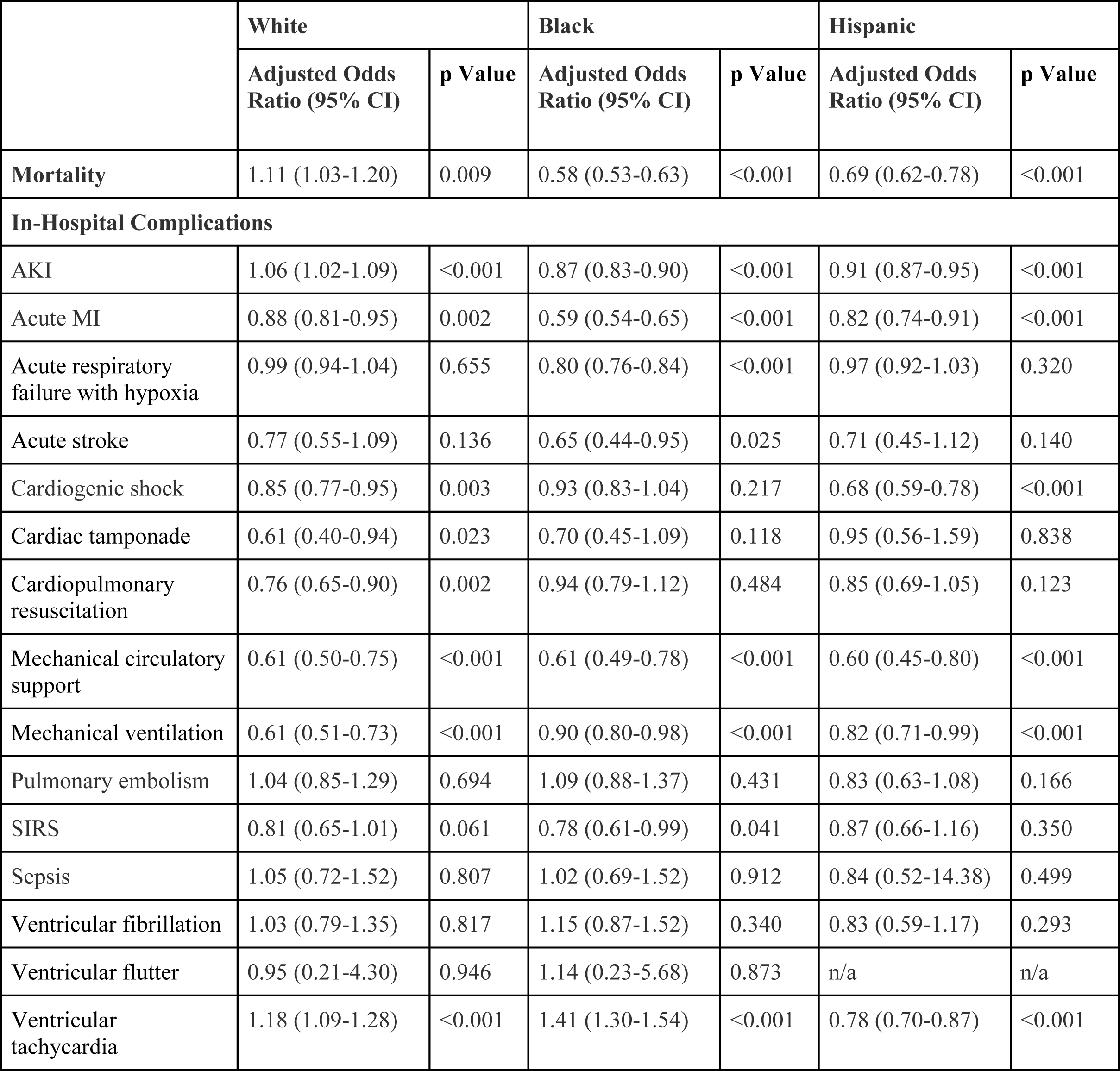
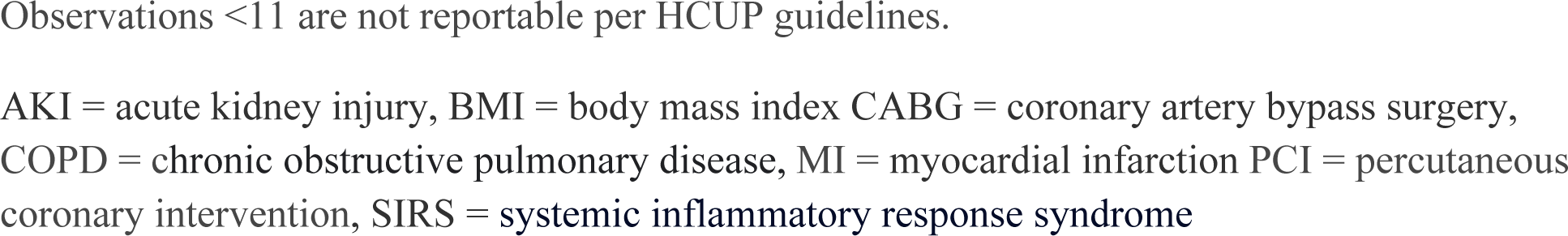
Adjusted Mortality and In-Hospital Complications Associated with HF and Advanced CKD Compared to Asian Patients.

### Income based hospitalized outcomes compared to Asian patients

White patients had significantly highest mortality across all four income quartiles after adjusting for comorbidities and gender. In contrast, Black patients were associated with lower odds of mortality compared to Asian patients across all income quartiles. However, Hispanic patients were only associated with significantly lower odds in the first quartile (OR: 0.69; 95% CI 0.61 to 0.79, p<0.001) and second quartile (OR: 0.64; 95% CI 0.54 to 0.76; p<0.001). Further details can be found in **Table 4**.

**Table 4.** Ethnic Differences in In-Hospital Mortality Compared to Asians Stratified by Median Income.

| **Income quartile** | **Adjusted Odds Ratio (95% Confidence Interval)** | **p Value** |
| --- | --- | --- |
| **1st Quartile (0-25th percentile)** |  |  |
| White | 1.11 (1.04-1.17) | <0.001 |
| Black | 0.57 (0.53-0.61) | <0.001 |
| Hispanic | 0.69 (0.61-0.79) | <0.001 |
| **2nd Quartile (26-50th)** |  |  |
| White | 1.17 (1.11-1.24) | <0.001 |
| Black | 0.64 (0.57-0.73) | <0.011 |
| Hispanic | 0.64 (0.54-0.76) | <0.001 |
| **3rd Quartile (51-75th percentile)** |  |  |
| White | 1.18 (1.11-1.25) | <0.001 |
| Black | 0.70 (0.61-0.81) | <0.001 |
| Hispanic | 0.89 (0.75-1.05) | 0.175 |
| **4th Quartile (76-100th percentile)** |  |  |
| White | 1.39 (1.30-1.47) | <0.001 |
| Black | 0.69 (0.57-0.83) | <0.001 |
| Hispanic | 0.90 (0.73-1.13) | 0.373 |

Observations <11 are not reportable per HCUP guidelines.

#### White compared to Asian patients in-hospital complications

In the first (OR: 0.81; 95% CI 0.67 to 0.99; p=0.045), second (OR 0.78; 95% CI 0.64 to 0.94; p<0.001), and fourth (OR: 0.91; 95% CI 0.83 to 0.97; p<0.013) income quartile, White patients had lower odds of requiring mechanical support. In the second (OR 0.75; 95% CI 0.64 to 0.86; p<0.001), third (OR 0.69; 95% CI 0.59 to 0.82; p<0.001), and fourth (OR: 0.83; 95% CI 0.70 to 0.99; p<0.046) income quartile, White patients lower odds of requiring mechanical ventilation.

#### Non-Asian compared to Asian ethnic patients in-hospital complications

In the first income quartile, Black patients had higher odds of mechanical ventilation (OR 1.38; 95% CI 1.22 to 1.56; p<0.001) and mechanical circulatory support (OR: 1.29; 95% CI 1.06 to 1.49; p<0.001). Similarly Hispanic patients were associated with higher odds of mechanical ventilation (OR 1.45; 95% CI 1.16 to 1.79; < 0.001) and mechanical circulatory support (OR: 1.45; 95% CI 1.16 to 1.79; p<0.001) in the first quartile. Hispanic and Black patients were similarly associated with higher odds of mechanical ventilation and mechanical circulatory support in the second quartile while no differences were seen in the third quartile. However, Hispanic were associated with lower odds of mechanical ventilation (OR 0.76; 95% CI 0.66 to 0.90; p<0.001) and mechanical circulatory support (OR: 0.80; 95% CI 0.72 to 1.04; p<0.001) in the fourth quartile. Black patients were also associated with lower odds of mechanical circulatory support (OR 0.88; 95% CI 0.76 to 0.98; < 0.001)

### Temporal trends

From 2016 to 2019, Asian patients with HF and advanced CKD decreased from 5,904 to 4,744. This compares to 32,501 to 36,139 in White, 14,222 to 17,708 in Black, and 5,470 to 6,556 in Hispanic patients. In 2020, trends in relative prevalence between the ethnic groups remained similar (**Figure 2**).

**Figure 2.**
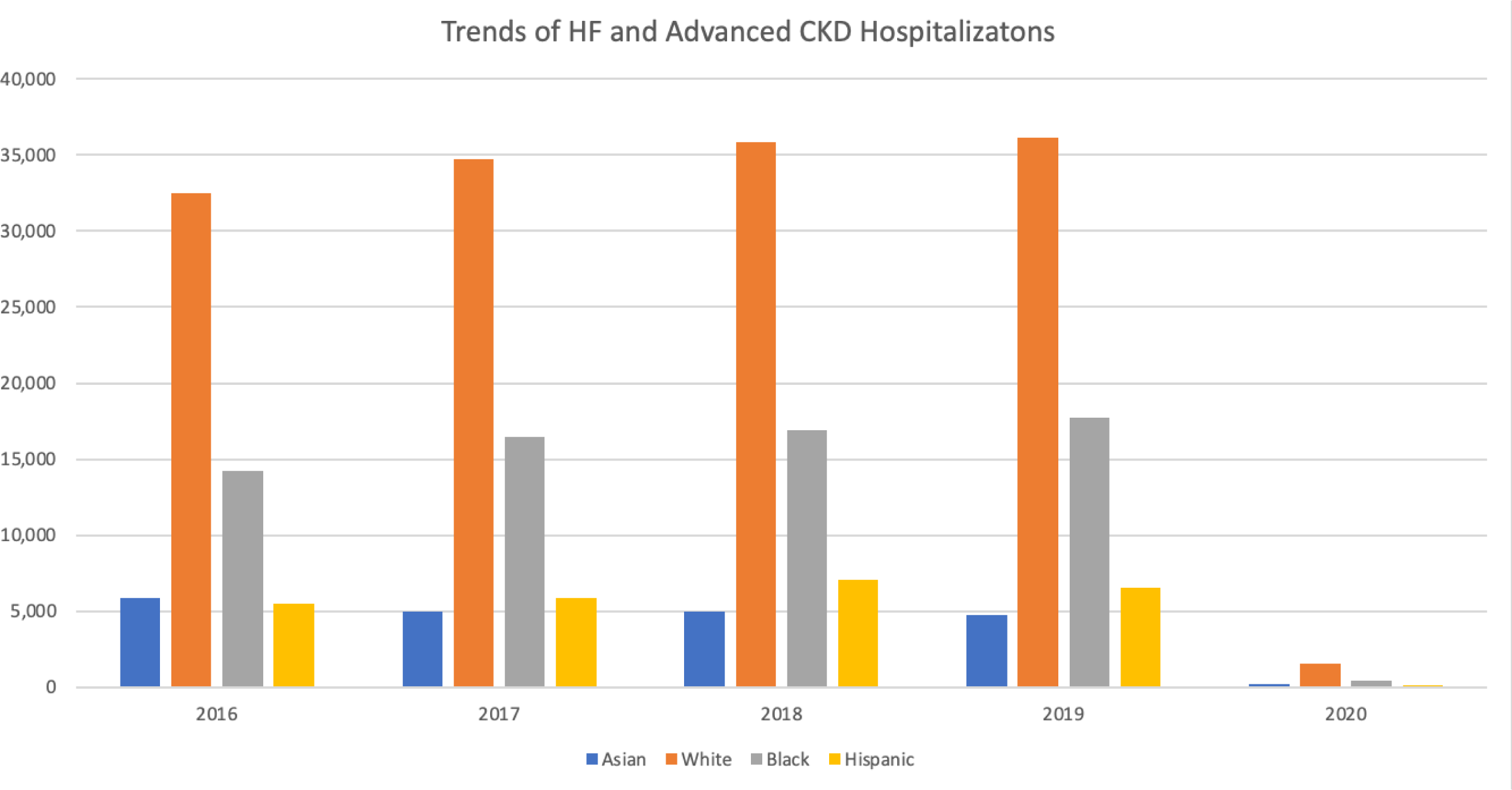
Prevalence Trends of HF Hospitalizations and Advanced CKD. Prevalence of Asian patients decreased between the study years. This is in contrast to increasing hospitalizations for all non-Asian ethnicities. Odds ratios were adjusted for Elixhauser comorbidity index, age, and sec using Asian patients as reference.

Over the study period, Asian patients had an increasing prevalence of cardiovascular comorbidities and in-hospital complications, including AKI, acute respiratory failure with hypoxia, sepsis, cardiogenic shock, mechanical circulatory support, and ventricular tachycardia (**Supplement Table 4**). Despite also increasing prevalence of White and Hispanic patients with HF and advanced CKD, their odds of in-hospital mortality decreased compared to Asian patients over the study period. In contrast, risk of in-hospital mortality remained relatively unchanged in Black and Asian patients (**Figure 3**).

**Figure 3.**
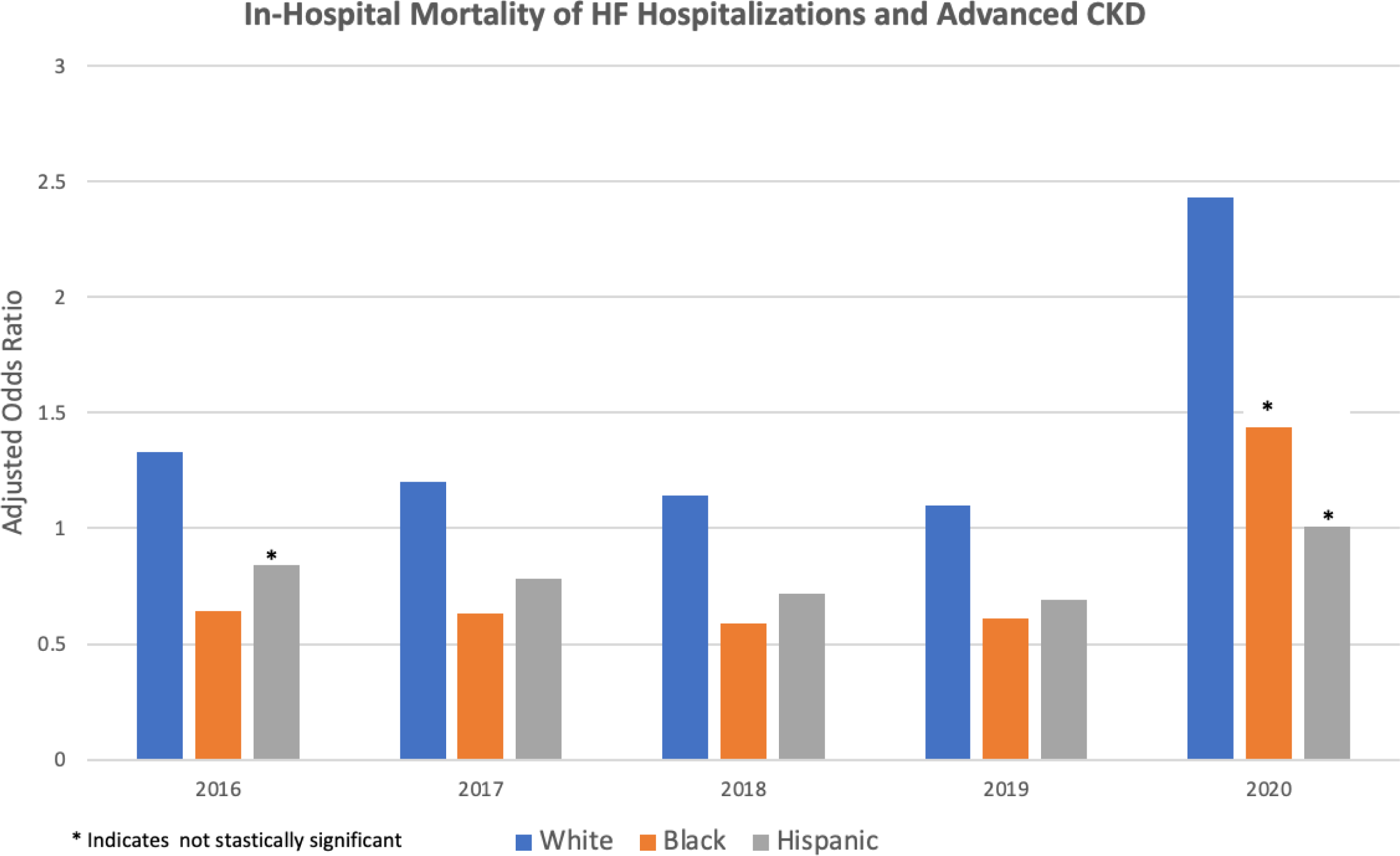
Odds Ratio for In-Hospital Mortality of HF Hospitalizations and Advanced CKD. Odds ratios for in-hospital mortality were higher for White patients and lower for Black and Hispanic patients. Odds ratios were adjusted for Elixhauser comorbidity index, age, and sec using Asian patients as reference.

### Length and cost of stay

Compared to Asian patients, White and Black patients on average had a lower hospitalization cost of $14,352.05 and $11.238.50 less but had an average longer duration of stay of 0.43 and 0.39 days, respectively (p<0.001). Compared to Asian patients, Hispanic patients had a hospitalization cost and length of stay on average $6,696 greater and 0.42 days shorter (**Supplement Table 5**).

## Discussion

In this study of 251,578 hospitalizations, we identified trends, compared outcomes and explored racial, ethnic, and socioeconomic disparities experienced by the Asian population with advanced CKD, who were hospitalized for heart failure exacerbation. A consistent decline in hospital admissions for Asian patients HF with advanced CKD was observed between 2016-2020. This trend stands in contrast to the growing prevalence of such admissions among non-Asian patients. There wa sa notable rise in prevalence of comorbidities including ischemic heart disease, valvular heart disease, primary and secondary preventative devices including ICD and pacemaker placement, compared to all other patients. However, higher prevalence did not correspond to equally high mortality or in-hospital complications, underscoring the existence of inequities in healthcare.

Non-Asian patients saw a decrease in overall adjusted complications across all income quartiles. Overall adjusted complications and costs related to HF and advanced CKD exhibited disparities based on race or ethnicity, as well as socioeconomic position. In our study, it was observed that Asian, Black, and Hispanic individuals exhibited the lowest median income. Furthermore, a higher percentage of Black and Hispanic patients were found to be situated among the two lowest income quartiles. This observation is also situated within the context of a higher representation of Black and Hispanic patients with Medicaid, which provides further evidence for the disadvantaged socioeconomic position of non-Asian ethnic patients. The distribution of median income exhibited heterogeneity, however, a general trend of decreasing prevalence was observed with higher income quartiles. Moreover, when compared to Asian patients, Black and Hispanic patients exhibited a lower odds of mortality across all median income quartiles. White and Black patients were noted to be associated with significantly lower hospitalization costs compared to Asian patients despite having a longer length of stay. This is in contrast to Hispanic patients who were associated with shorter length of stay and lower hospitalization costs, compared to Asian patients. The comorbidity burden of White patients had significantly higher comorbidity burden which is reflected in the higher odds of mortality. While Hispanic and Black patients also had higher comorbidity burden compared to Asian patients, the difference was less suggesting other factors may be contributing to difference in mortality and complications. more evenly distributed median income and reduced comorbidity burden, Asian patients continue to experience considerable healthcare disparities in contemporary times. These disparities manifest in greater hospital costs, longer hospital stays, increased mortality and in-hospital complications.

Heart failure is a complex clinical syndrome where there is an impairment of adequate distribution of blood in the systemic circulation. In advanced CKD, patients are more susceptible to fluid overload, raised blood pressure due to endothelial dysfunction, arrhythmogenic and atherosclerotic disease. In recent years, there has been a consistent and stable prevalence of HF among the elderly demographic, alongside the presence of risk factors commonly associated with heart failure, such as type 2 diabetes, obesity, and renal failure.^10,11^ Research has shown that a significant proportion, ranging from 50% to 70%, of patients admitted to the hospital for HF exhibit the presence of at least one non-cardiovascular risk factor.^12^ Among the many risk factors, it was found that CKD had a prognostic consequence of 41% on death and rehospitalization.^13,14^ It is widely recognized that individuals admitted for HF commonly belong to the lowest quartile of income and are frequently from ethnic minority groups.^15^ The findings of our study corroborate a comparable pattern observed in hospitalized patients diagnosed with HF and advanced CKD.

Our study has shown significant disparities faced by Asian patients which may contribute to variations in outcomes. Certain subgroups within the Asian population have distinct genetic mutations, such as higher frequencies of CYP2C19 alleles and UMOD, that have an impact on their response to medical intervention^16,17^ Various genetic markers have demonstrated differential effects on glomerular filtration rates across racial groups, influencing CKD progression and function, and ultimately impacting treatment outcomes.^18^ Additional affected regions encompass varying reactions to pharmaceuticals such as ACE-inhibitors or diuretics, as well as other genetic indicators that predispose the Asian demographic to cardiorenal syndrome.^19^

Effects of culture, communication, and language barrier specific to nonnative English speaking patients, cultural differences and practices, and social ties are well documented in literature as an under addressed cause of healthcare disparity. HF exacerbation and advanced CKD are complex clinical situations that necessitate effective communication and a shared understanding between patients and healthcare providers. In this context, language and cultural competency are critical. Research has identified several prominent themes that emerge in the provision of healthcare to patients of Asian descent. These themes encompass the significance of cultural knowledge, the influence of familial networks, the impact of dietary modifications, and the relevance of traditional beliefs. This barrier is especially more consequential in acute and complex patients, which often necessitate the involvement of a diverse, large interdisciplinary team for management, such as individuals with HF and advanced CKD.^20–22^ Cultural beliefs often lead to incongruent perceptions of disease severity, causing delays in seeking medical attention and even declining care. This issue is compounded by language and communication obstacles, resulting in incomplete patient evaluations, delayed treatment, limited grasp of the patient’s condition and recommended therapies, and a reduced trust in the quality of care they received.^23,24^ Many patient-physician interactions involve the utilization of web translating services or impromptu, untrained interpreters, despite existing evidence underscoring the hazards linked to these practices.^25^ This possibility of linguistic and cultural competency as a contributing factor in outcomes of Asian with advanced CKD admitted for HF is further supported by our study. Specifically, our study revealed no significant difference in complications in the third income quartile, and a decreased association of complications in the fourth income quartile, and overall lower mortality in the Black and Hispanic population compared to Asian patients. This is significant because higher income is associated with greater and more equal access to healthcare across race/ethnic group, healthcare literacy, and education yet we report differences in outcomes.^26^

English is the primary language spoken in the United States, followed by Spanish. In the United States, there exists a significant difference of nearly ten times in the number of individuals proficient in Spanish compared to those proficient in any Asian language.^27^ It is well documented that addressing language and competency barriers can significantly enhance healthcare outcomes for Asian patients. This is particularly important in HF in the setting of advanced CKD, a complex and largely clinically treated disease that depends on clear communication to understand, treat, and prevent complications. Multiple studies demonstrated supporting the efficacy of multilingual healthcare setups and culturally tailored intervention in improving clinical care and healthcare outcomes.^28,29^ Furthermore, research has demonstrated that racial concordance, which frequently coincides with a shared cultural understanding, plays a significant role in fostering a positive patient-physician relationship and ultimately leading to better healthcare outcomes.^30^ Nevertheless, there continues to be an underutilization of professional interpreters in the healthcare setting. In one study, 57% of residents reported usually or always using a professional interpreter despite 97% endorsing the advantages associated with enhanced outcomes..^31^ Access to these services also varied with only 68% of US hospitals offering language assistance services, including both remote and in-person services, while 27.9% of patients reported that no interpreter was accessible when needed during hospitalization.^32,33^ Lastly, a study by the Joint Commission found that only 13% of hospitals successfully met all four standards pertaining to language-related culturally and linguistically appropriate services, whereas 19% met none.^34^ These findings highlight the need of addressing these disparities through implementation of policies that encourage investments social risk assessment tools that are sensitive to Asian and other ethnic populations. Additionally, it is crucial to develop solutions that enable healthcare providers to effectively address the needs of socially vulnerable patients.

### Strength and limitations

The NIS database aggregates data from millions of hospital stays annually and enables for powerful statistical analysis of special patient populations throughout multiple years. Furthermore, we conducted an analysis of long-term trends, providing valuable insights for policymakers and other pertinent stakeholders. The utilization of data derived from both community and academic institutions contributes to an enhanced comprehension of healthcare patterns and results.

However, these strengths are accompanied by inherent limitations of the NIS database. First the available data does not provide sufficient information regarding outpatient settings or post-discharge care. Furthermore, the collection of readmission rates and prior admission data are not captured. Since readmission is common in HF, our study is unable to differentiate the presenting severity that justified hospitalization. This limitation leaves a gap in the continuum of care where the root cause and other contributors to outcomes can be missed. Additionally, the inability to disaggregate ethnic groups contributes to significant heterogeneity. Prior studies have demonstrated differences in cardiovascular outcomes in the three predominant Hispanic groups (Mexican, Puerto Rican, Cuban individuals) in the United States.^36^ Other limitations include coding errors and inaccuracies, which can inadvertently introduce biases in the research. Moreover, the lack of granularity in the data that precludes nuanced analysis such as laboratory values, imaging duration, and treatment regimen of HF. This information would be beneficial in exploring the impact of language and culture barriers in care of advanced CKD patients admitted for HF through other surrogate markers.

## Conclusion

In summary, we present findings that demonstrate notable disparities in in-hospital outcomes and complications among Asian patients with advanced CKD admitted for heart failure. More qualitative research is needed to evaluate and supplement our findings to identify how clinical practices can better incorporate language and cultural understanding into heart failure and CKD practices and close the gap in complications, mortality, and cost and length of stay. Further investigation would also be beneficial to examine whether advanced CKD patients belonging to specific ethnic or racial groups experience reduced negative outcomes during hospitalizations for HF when they are enrolled in services provided by dedicated specialized multidisciplinary medical teams.

## Data Availability

Publically available data

## Non-standard Abbreviations and Acronyms

ECMO: Extracorporeal membrane oxygenation
HF: Heart failure
IABP: Intra-aortic balloon pump
pVAD: Percutaneous ventricular assist device
SIRS: Systemic inflammatory response syndrome

---

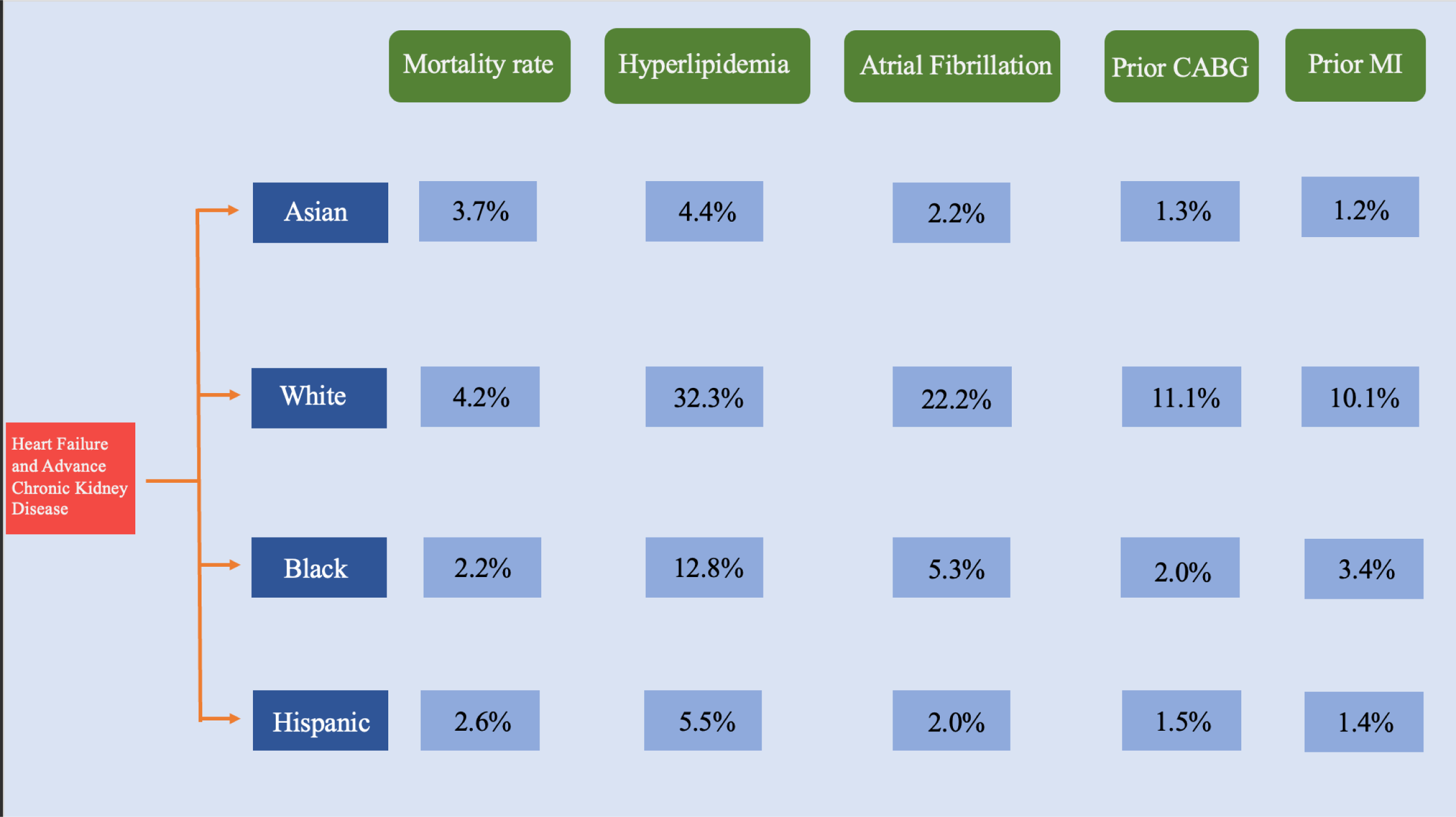
Central Illustration. Racial Disparities in Clinical Comorbidities of Patients with Heart Failure and Advanced Chronic Kidney Disease. White, Black, and Hispanic patients have a greater prevalence of cardiovascular comorbidities disease compared to Asian patients. However, Black and Hispanic patients have a lower mortality rate.

